# A multi-country qualitative study among hospital patients and carers perspectives on living with multimorbidity: a case of Malawi and Tanzania

**DOI:** 10.1101/2025.10.27.25336133

**Authors:** Sangwani Salimu, Stephen A. Spencer, Alice Rutta, Treighcy Gift Banda, Ibrahim Simiyu, Nateiya M. Yongolo, Graciana Kimario, Gimbo Hyuha, Martha Oshoseny, Marlen Chawani, Augustine Choko, Paul Dark, Julian T. Hertz, Blandina T. Mmbaga, Juma Mfinanga, Rhona Mijumbi, Adamson S. Muula, Mulinda Nyirenda, Francis Sakita, Charity Salima, Hendry Sawe, Miriam Taegtmeyer, Jamie Rylance, Eve Worrall, Felix Limbani, Nicola Desmond, Deborah Nyirenda, Ben Morton, MultiLink Consortium

## Abstract

**Background:** Multimorbidity is an urgent public health threat in sub-Saharan Africa (SSA). However, data on the experiences of people living with multimorbidity (PLWMM) in this context are limited. We explored patient and carer experiences of living with (or caring for) multimorbidity to inform the development of patient-centred interventions for managing multimorbidity.

**Methods:** This qualitative study is nested within a broader programme of multimorbidity research conducted in Malawi and Tanzania across four hospitals. We recruited patients recently discharged from hospital with known two or more combinations of hypertension (HTN), diabetes mellitus (DM), HIV and chronic kidney disease (CKD) and their carers. We conducted primary in-depth interviews at discharge and follow-up interviews 90 days after initial hospital admission to explore longitudinal experiences and care trajectories. FGDs were conducted after hospital discharge. Data were analysed thematically and presented through the lens of an existing Expanded Conceptual Model on Multimorbidity for SSA.

**Results:** We conducted 32 in-depth-interviews (IDI) and 8 focus group discussion (FGDs) with PLWMM and carers in Malawi; and 21 IDIs and 7 FGDs in Tanzania. We identified, and present findings under three key crosscutting themes: experiences of living with multimorbidity; self-management and adaptation; and prioritisation of individual diseases within the multimorbidity paradigm. Age, sex, disease combinations and settings impacted on experiences living with multimorbidity. Out-of-pocket expenditure and poor quality of care dominated both settings with CKD and DM comorbidities exerting the heaviest burden on PLWMM and carers. Treatment discontinuation was common for HTN in Malawi and CKD in Tanzania, whilst living with HTN was linked to emotional distress in both. Older PLWMM reported greater family disruption due to loss of independence. Health crises, health literacy, and financial constraints were major drivers of disease management. Individuals particularly experienced stigma when conditions caused visible signs, and described moral and spiritual concerns.

**Conclusions:** Multimorbidity experiences in Malawi and Tanzania reflect complex interactions between individual, socioeconomic, and health system factors. Effective interventions require multidisciplinary, patient-centred approaches addressing structural barriers, improving health literacy, and promoting collaborative care involving patients, carers, and peers.

## Introduction

The World Health Organization defines multimorbidity as the coexistence of two or more chronic conditions (1). Multimorbidity is associated with negative health outcomes (2), and presents a complex and growing challenge in sub-Saharan Africa (SSA) (3, 4), increasing pressure on already strained healthcare systems (5). The interaction of chronic communicable (CDs) and non-communicable diseases (NCDs) is particularly problematic in SSA where HIV prevalence remains high. Researchers increasingly recognise the potential catastrophic impacts of multimorbidity in SSA but, to date, there has been inadequate exploration of patient and carer perspectives (6).

Despite increasing recognition of the burden of multimorbidity in SSA, patient-centred care remains underdeveloped. Our recent systematic review and meta-synthesis found a paucity of data on self-management of multimorbidity among patients and their carers in SSA (7). Key challenges include poor existing health literacy; limited support to increase health literacy; low self-efficacy among PLWMM and carers; and under developed health services as barriers to effective self-management. The systematic review highlights the need to strengthen self-efficacy among PLWMM and their carers, to enable better control of their conditions; improve treatment adherence; and make necessary lifestyle adjustments (8). For example, in South Africa (9, 10), Tanzania (11) and Malawi (12) PLWMM and their carers often struggle to balance complex ill-health with treatment requirements and managing complications of the conditions. Poor health literacy negatively impacts the ability to understand and manage conditions (11, 13). These issues are frequently compounded by limited access to care, which may be fragmented and difficult to navigate (10, 12, 14, 15).

The aim of this study was to explore patient and carer experiences of living with multimorbidity (or caring for) PLWMM in Malawi and Tanzania to inform the development of patient-centred interventions, improve management and reduce the risk of complications (3). We sought to understand how PLWMM and their carers prioritise disease management using in-depth interview (IDI) and focus group discussions (FGDs). Generation of this patient-centred data is vital to inform context-relevant approaches designed to effectively and sustainably improve health outcomes in the SSA region.

## Methods

### Design

We conducted a qualitative longitudinal study using IDIs and FGDs using an iterative approach to data collection. We incorporated intercountry comparisons to explore contextual similarities and differences across settings (16). Insights from early IDIs and FGDs were used to guide and refine subsequent sessions, to enable a deeper understanding of the issues. We collected demographic information including age, sex, marital status, education, conditions and residence. We used the Consolidated Criteria for Reporting Qualitative Research (COREQ) (17) to present findings (see supplementary materials [Table S1].

### Setting and context

The study was conducted in two settings, Tanzania, a low-middle income (LMIC) and Malawi, low-income country (LIC) (18, 19) respectively. Both contexts are subject to rapid shifts in demographic and disease patterns, characterised by ageing populations; increased prevalence of non-communicable diseases (NCDs) (20, 21); and strained health systems (18, 19). We recruited study particpants at four facilities, two referral and two district hospitals, Queen Elizabeth Central Hospital (1,350 bed capacity), and Chiradzulu District Hospital (300 bed capacity) in Malawi; and Muhimbili National Hospital (1,500 bed capacity), Dar-es-Salaam and Hai District Hospital (140 bed capacity) in Tanzania. Public healthcare services are provided free of charge in Malawi (22) while Tanzania has adopted a cost-sharing and health insurance model (23, 24). Persistent bottlenecks across both these health systems frequently lead to catastrophic out-of-pocket (OOP) health expenditures. The success of HIV interventions, primarily funded by PEPFAR and the Global Fund, has led to improved life expectancy (25, 26). However, the concurrent rise in NCDs remains underfunded, with limited resources dedicated to addressing this emerging health challenge (27, 28). The study settings are further described in our overarching study protocol (3) and results (29).

### Sampling and recruitment of participants

We used a purposive recruitment strategy to identify PLWMM and their carers already recruited to our overarching prospective cohort study (3). Research nurses approached cohort study participants for additional recruitment to our qualitative study (30). Researchers for this sub-study had no prior relationships with participants. Participants were included if they were recruited to the overarching study; aged ≥18; recently discharged from one of four hospital facilities; aware of their multimorbidity diagnosis (chronic kidney disease [CKD], diabetes mellitus [DM], hypertension [HTN] and/or HIV); capable of providing informed consent; had capacity to participate in interviews or discussions; and lived within defined facility catchment areas. Carers were included if they were aged ≥18 and provided regular care to a PLWMM recruited to the overarching study. We excluded PLWMM or carers under 18 or who were unable to provide written consent. We deliberately did not pair patients with carers for this study to explore broader experiences while avoiding potential bias or repetition in responses. We categorised the data by country, age, gender, and disease combinations to further inform our findings.

### Data collection

Data collection was facilitated by SS (a PhD student) in Malawi and IS, GK, MO, and AR in Tanzania - all trained social scientists with experience in qualitative methods. Relevant tools were translated into local languages (Chichewa in Malawi and Swahili in Tanzania). All participants and facilitators were fluent in the respective local language. Data collection took place from 10/01/2023 to 20/10/2023, in both contexts, with interviews conducted at discharge and again 90 days after initial hospital discharge. The 90-day interviews were planned to coincide with planned medical follow-up visits to the health care facility where possible. Follow-up interviews aimed to explore how care continued after hospital discharge (home and community), and during follow-up hospital visits. We scheduled FGDs at the point of hospital discharge and met at centrally located venues, convenient for participants. SS, from Malawi, developed the IDI guide, and collaborated with the Tanzanian teams to coordinate refinement and standardisation. Drawing on principles from qualitative research networks (QRNs), we used harmonised tools adapted to local contexts (16, 31). These were then piloted in both settings to identify and address ambiguities, cultural nuances, and contextual challenges. To maintain consistency across locations, data collectors from both sites participated in joint training and feedback sessions. We used data triangulation to leverage both individual and group-level insights to enhance our understanding (32, 33). IDIs provided detailed insights into lived experiences, including those of patients too ill to participate in FGDs. FGDs facilitated dynamic discussions, to enable participants to share experiences and enhance their collective understanding of living with multimorbidity (32). We conducted member checking by asking participants to confirm and refine summaries and interpretations of emerging themes at the end of each session (34, 35). Sessions were conducted until thematic saturation was reached, to ensure both consensus and representation of diverse viewpoints (36, 37).

We recruited 185 participants across both settings and conducted 15 FGDs and 127 IDIs (initial and follow-up). Of these, 89 were patients and 96 were carers. In Malawi, 94 participants (50 females, 44 males) took part in 8 FGDs and 56 IDIs (32 initial, 24 follow-up), while in Tanzania, 91 participants (48 females, 43 males) participated in 7 FGDs and 71 IDIs (41 initial, 30 follow-up) (Figure 1). The initial interviews in both settings were split equally between PLWMM and carers. FGDs were stratified by gender. IDIs lasted between 45 minutes to 1 hour while FGDs lasted 1 hour to 1 hour 15 minutes. In Malawi, FGDs were equally split between males and females, while in Tanzania, there were more male FGDs (4) than female (3). Only study facilitators attended IDIs, but FGDs were supported by research nurses who obtained informed consent from participants. Additionally, the number of follow-up interviews was reduced due to death or serious illness of some PLWMM participants. Follow-up interviews were not conducted with carers of deceased patients to avoid further distress.

**Fig 1.**
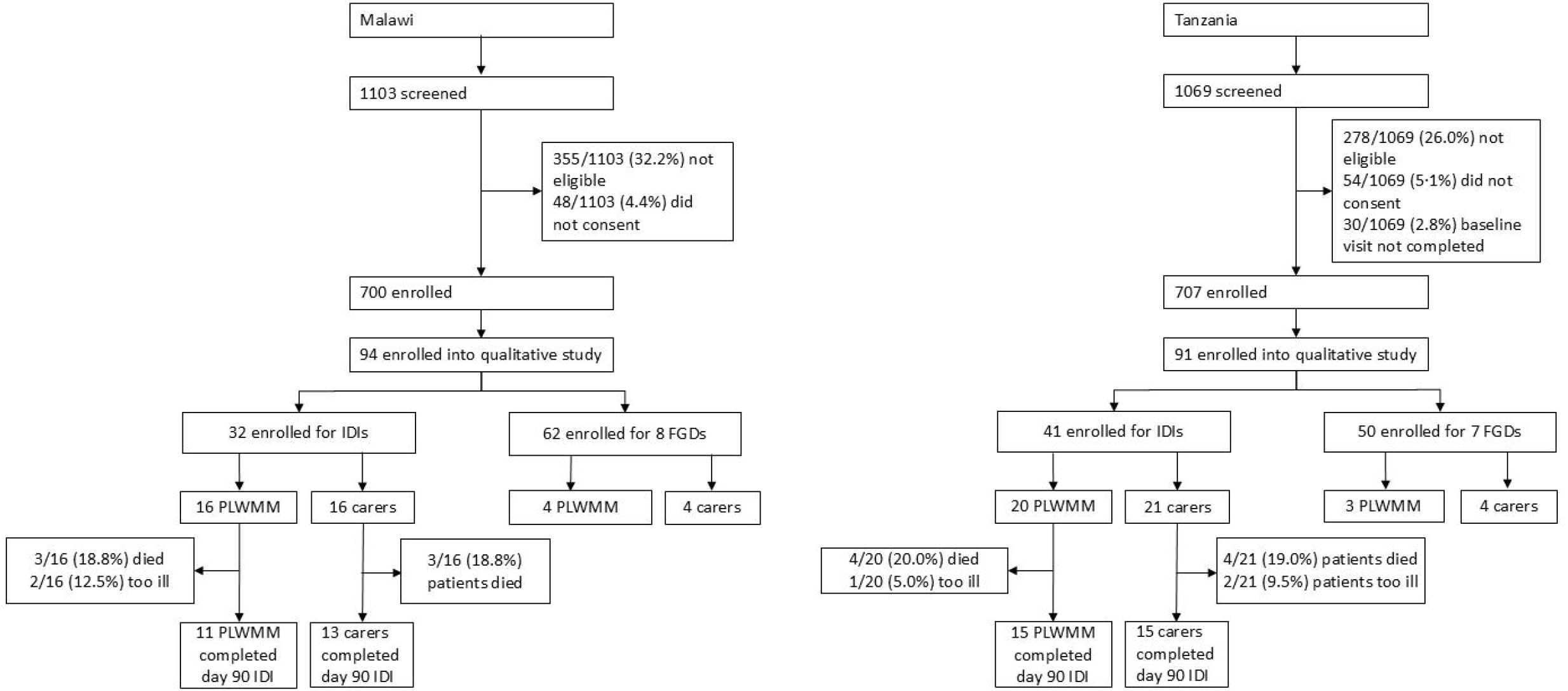

### Data management and analysis

All sessions were recorded, transcribed verbatim, and anonymised. Transcripts were translated into English. Data management and analysis were conducted using NVivo 12 Pro, (QSR International, Melbourne, Australia, released 2023). We used thematic analysis, following Braun and Clarke’s six-step approach, to identify patterns and meanings within the data (38). SS coded all data with initial familiarisation by repeatedly reading the transcripts and documenting initial impressions. Next, an inductive approach was used to generate initial codes and systematically identify meaningful segments of text. These codes were then grouped into potential themes, reflecting key aspects of participant experiences and perspectives. The themes were iteratively reviewed and refined in collaboration with the research team to ensure they accurately represented the dataset and captured the diversity of participant experiences. Each theme was clearly defined, and representative quotes were selected to illustrate key findings. The final themes were synthesised to provide a deeper understanding of multimorbidity experiences; access to healthcare; and potential interventions to improve patient outcomes. To facilitate cross-country comparability, each country team conducted initial analyses independently, followed by joint virtual meetings to compare and refine emerging themes. This collaborative approach facilitated the identification of both convergent and divergent themes, enhancing the analytical rigor and contextual relevance of the findings (34, 35). This methodology aligns with practices observed in multi-country qualitative research consortia, such as the STAR and REACHOUT projects, which emphasize iterative, collaborative analysis and reflexivity to ensure methodological consistency across diverse settings (16). No single condition (HIV, DM, HTN or CKD) was treated as a standalone analytic category. Instead, the analysis centred on the lived experience of multimorbidity as a whole. However, we paid close attention to variations by age, sex, geographic location, and combinations of conditions, as these factors often shaped participant care experiences; illness narratives; and interaction with the health system. This allowed us to capture important social and structural patterns without fragmenting the data by disease.

### The Expanded Conceptual Model on Multimorbidity

We applied the Expanded Conceptual Model on Multimorbidity (6) post-analysis to structure and interpret our findings; assessing how well the identified qualitative themes aligned with its theoretical premises. The model suggests a shift from a disease-counting approach towards a more nuanced understanding of common co-occurring health conditions. The suggested approach foregrounds cascading burdens experienced by individuals, families, communities, and health systems, whilst also identifying factors such as meaningful improvements in social or health support that can reverse or mitigate decline. The model also emphasises the importance of both primary and secondary prevention of disease and shifts attention from biomedical and, behavioural interventions to interdependent care structures rooted in social and systemic change; inter- and, transdisciplinary collaboration and participatory engagement with PLWMM and, their communities. The tool integrates syndemic frameworks (40), which examine the interconnection of health issues within social, economic, and cultural contexts, and complexity theory (41), emphasising a contextual understanding of patient environments and needs. Additionally, Burden of Treatment Theory (12, 42) provided insights into the relational dynamics of patient workload, framing patients as active agents influenced by social networks and healthcare organisation. The strength of the model lies in its integration of multiple theoretical perspectives and its holistic, person-centred orientation. By mapping our emerging themes onto this framework, we aimed to evaluate its relevance and applicability in the SSA context, particularly given the limited existing evidence on multimorbidity in the region (12, 39).

### Reflexivity and positionality

SS has direct experience of the public healthcare system in Malawi and led data collection in this setting, her role in Tanzania was limited to training data collectors and remote oversight of research processes. Acknowledging this, SS actively engaged with the Tanzanian research team to incorporate local insights and refine the analysis. For example, a key difference was government supported transport services for patients in Tanzania (not in Malawi), provided at a cheaper rate than private providers. To enhance credibility and generalisability, SS collaborated with senior co-authors and study team members, leveraging their diverse expertise. Local researchers (IS, AR) and data collectors (IS, GK, MO) who are fluent in Swahili and familiar with local norms and healthcare settings, played a crucial role in contextualising participant narratives. Member checking and regular team reflections supported confidence in the data interpretation and contributed to a rigorous, collaborative analytical process.

Given the potential for financial incentives or perceived healthcare benefits to influence participation (43), trial nurses clearly explained study independence from clinical services and reinforced that participation was voluntary. To build rapport and minimise social barriers, all data collectors explicitly identified themselves as social scientists rather than healthcare providers. Researchers were reflexive about power dynamics throughout, continuously adapting approaches to ensure an accurate and respectful representation of participant perspectives.

### Ethical Statement

Ethical approval was obtained from LSTM (21-086); College of Medicine Research and Ethics Committee (COMREC), Malawi (P.11/21/3462); National Institute for Medical Research (NIMR), Tanzania (NIMR/HQ/R.8a/Vol.IX/4008; Kilimanjaro Christian Medical Centre (KCMC) (2570). We obtained written (or witnessed thumbprint if the participant was illiterate) informed consent from all the participants.

## Results

### Participant descriptions

The median age for PLWMM in IDI in Malawi was 50 (IQR 44–57, range 26–69) while in Tanzania the median age was 60 (IQR 47-66, range 31-80). For carers, the median age for in IDI in Malawi was 55 (IQR 39-58, range 22-71) while in Tanzania the median age was 53 (IQR 34-61, range 24-73). The most common disease clusters among PLWMM included: DM and HTN (10 patients); HIV and HTN (8 patients); HTN, DM and CKD (4 patients) represented the most common triple combination (see Figure 2). Heart failure (6 patients) and TB (5 patients) were the most common conditions identified outside the focus diseases. These combined figures reflect data from both Malawi and Tanzania. Details for PLWMM engaged in IDIs are presented in Tables S1 (Malawi) and S2 (Tanzania).

**Fig 2.**
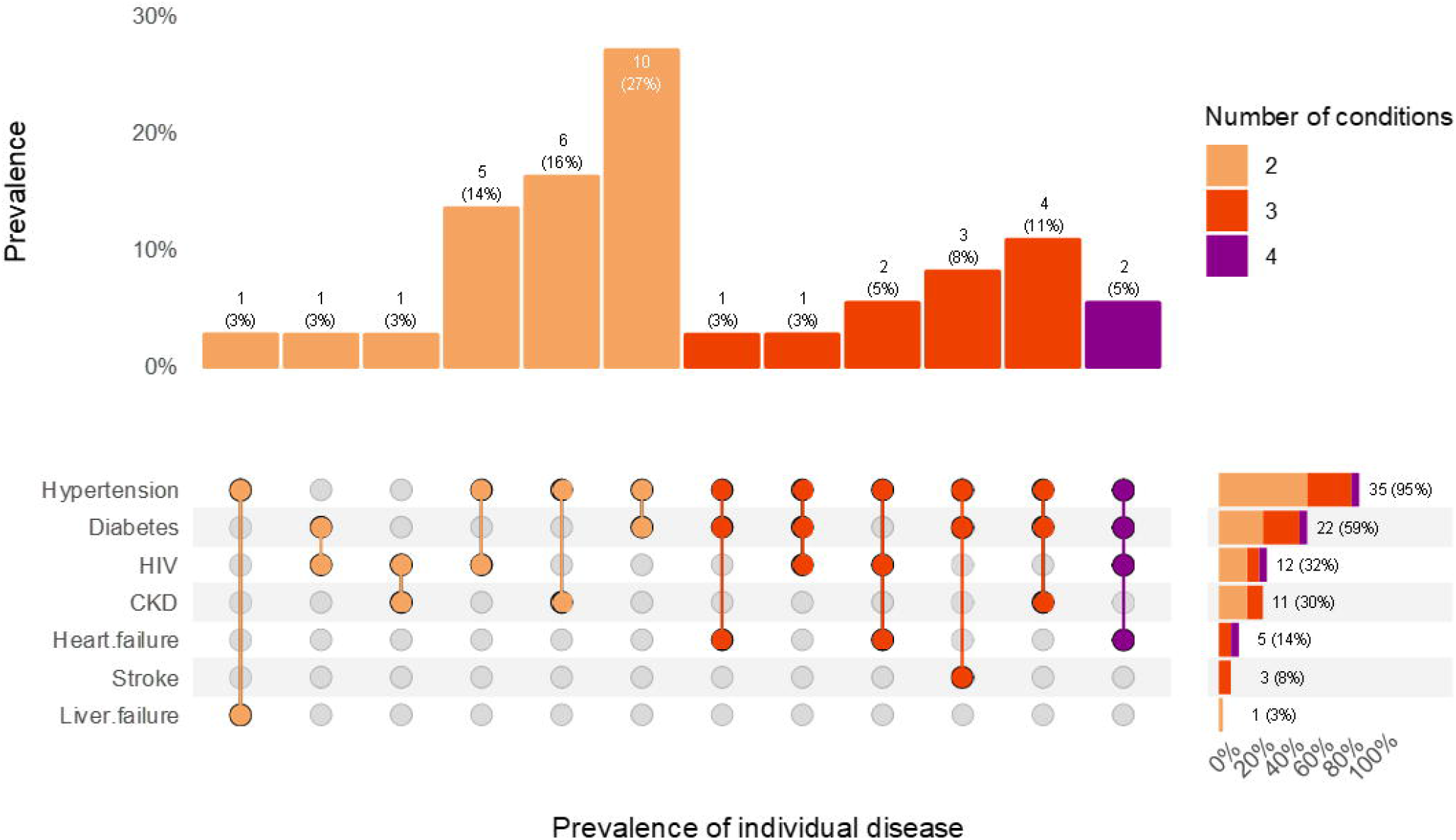

We present findings under three themes: experiences of living with multimorbidity, self-management and adaptation; and prioritisation of individual diseases within the multimorbidity paradigm (Table 1). These similarities and differences between perspectives in Malawi and Tanzania are explored in each theme.

**Table 1:** Themes.

| Theme | Sub themes |
| --- | --- |
| Experiences of living with multimorbidity | Lack, loss and deprivation |
|  | Psychological, emotional disorders are common |
|  | Impact of lifestyle and environment |
|  | Support |
| Self-management and adaptation | Managing means balancing multifaceted realities |
|  | Reliance on biomedical treatment |
|  | Critical defining moment |
|  | Reduced use of alternative treatments |
| Prioritisation of individual diseases within the multimorbidity paradigm | Disease-centric approach among PLWMM is more common among HIV- than non- HIV cluster combinations |
|  | Temporary discontinuation of antihypertensive drugs |

### Experiences of living with multimorbidity

PLWMM and their carers described multimorbidity as a major disruption to their daily lives, affecting their physical health and psychological well-being; with broad impacts on social participation and interactions. Across both settings, PLWMM and carers described how multimorbidity caused reduced physical functional ability and restricted work capacity. This impacted on their ability to self-manage; with loss of financial independence and decreased social standing within the family. These in turn caused anxiety and depression amongst participants. These issues were particularly acute for people living with DM or CKD comorbidity in both settings. While disability increased with the number of conditions per individual, DM or CKD comorbidity were felt to be more stressful because of their physical complications.

> *“I feel unwell, weak, can’t walk long distances. I get tired easily. I don’t have any abdominal pain, but the bloating is very uncomfortable,”* Tanzania IDI PLWMM Male 30-39 (HTN, CKD).

Beyond the cumulative effect of living with multimorbidity, older PLWMM experienced greater physical limitations and loss of independence compared to their younger counterparts leading to an increased reliance on carers and family support. Disruption to daily life stemmed from the shifting balance between independence and reliance on others for daily care and support.

> *“Chronic illnesses disrupt families…. The expensive treatment affects the family’s economy because people think about the illness all the time*… *For me, they are constantly asking, ‘has mum taken her medicine?’,”* Tanzania IDI PLWMM Female 60-69 (HTN, DM).

Older PLWMM across both settings were more likely to describe how personal stressors impacted health outcomes, particularly with HTN. Emotional stress, arising from family conflict; financial strain; and the cumulative burden of caregiving and illness was not only seen as a response to their deteriorating health, but also perceived as a trigger for the onset or worsening of chronic conditions. Participants frequently noted how psychological distress directly impacted their physical health, reflecting the complex interplay between emotional well-being and disease progression. This perceived connection between stress and illness was especially evident in narratives where individuals attributed the emergence of hypertension or worsening symptoms to emotionally charged events.

> *“I argued with my parents…since then, I developed high blood pressure,”* Malawi IDI PLWMM Female 50-59 (HIV, DM, HTN).
>
> *“My son’s arrest and life imprisonment led to my first illness* (hypertension)*…. Then, my younger son died. Two years later, my wife died, … that’s when my problems began,”* Tanzania IDI PLWMM Male 70-79 (HTN, DM).

Women across both settings described how living with multimorbidity or caring for a patient was compounded by the simultaneous need to care for others. Their narratives reflected a dual burden of managing their own illness while attending to the health and daily needs of family members. This strained their time and resources; disrupted income-generating activities; and made individuals feel exhausted.

> *“It’s a big responsibility* (taking care of her sister with multimorbidity), *I hope I won’t get into trouble with my family…. They know my sister is sick but may not fully understand …. We need help but I am usually with her, so how can I make money? I am usually exhausted,”* Tanzania FGD Carer Female 40-49.
>
> *“A lot changed because he is ill…. He was a tailor but stopped…. now everything depends on me. I look after him. It takes a lot of time,”* Malawi FGD Carer Female 40-49.

Some participants described experiences of stigma and how they dealt with it. To mitigate the emotional toil of HIV-related stigma, some PLWMM reasoned through their experiences and reframed their diagnosis and treatment, casting HIV as a manageable condition. Participants linked stigma to the visible side effects of HIV or CKD treatments.

> *“My relatives would mock me, telling others ‘he is taking ARVs*.*’ That disappointed me. After reflecting, I noted that we do take drugs like Panadol, only that with ARVs, it’s daily, so I decided not to mind them because I would be stressed. Before testing for HIV, I would often get sick, would buy Panadol, be okay for two days and then fall sick again. When I started taking ARVs, my condition improved and that means ARVs help me to stay healthy,”* Malawi FGD PLWMM Male 40-49 (HIV, HTN).
>
> *“Caring for a patient who is stressed because of multiple diseases is difficult…. Even if the patient is positive minded, some people mock them because of how they look, but we must interact with them,”* Tanzania FGD Carer Female 60-69.
>
> In some instances, it was difficult to determine whether patient actions were motivated by self-stigma. However, the deliberate separation of care such as seeking ART in one facility while accessing DM and HTN services in another may suggest an internalised fear of being identified or judged, even in the absence of overt external stigma.
>
> *“I visit diabetes and hypertension, here* (tertiary facility), *ART in* (neighboring district hospital) where I was diagnosed. *Here* (diabetes clinic), *they just ask that do you go to thing… they just make sure I keep my appointments at the ART clinic,”* Malawi IDI PLWMM Female 50-59 (HIV, DM, HTN).

In addition to coping with stigma and the visible markers of multimorbidity, patient narratives illustrated how community perceptions of illness causality shaped their day-to-day lives. Common across both settings, complications from DM and CKD presented the most exposure to stigmatization due to visibility of disease.

> *“I also have Hepatitis B and hypertension…. The hepatitis is bad, my tummy is always swelling. It’s a heavy burden people assume I stole or was bewitched…. I stopped working, I am now like a disabled person or a beggar,”* Tanzania FGD PLWMM Male 40-49 (HIV, CKD).

Limited access to information contributed to uncertainty and confusion about diagnoses and treatment, particularly among patients living with CKD comorbidity in Malawi. Some individuals were newly diag-nosed and had not yet received a full explanation of their condition, while others expressed they were not aware of the treatment they were taking.

> *“I don’t know when the diagnosis was made, the time I knew, they just told me this medication is for BP, this medication is for kidneys,”* Malawi IDI PLWMM Male 50-59 (HTN, DM, CKD).

PLWMM and carers experienced frustration and fear related to the unpredictability and worsening of their conditions, which intensified feelings of helplessness. PLWMM expressed shock at the emergence of new chronic conditions. This was more common as the number of conditions per individual increased, indicative of progressive deterioration.

> *“The new one is worse than the previous. I started with heart problem, then diabetes and then kidney problem. With time, a new one comes and it affects you me more than the previous ones,”* Tanzania IDI PLWMM 50-59 Male (HTN, DM, CKD).
>
> *“I wonder, what’s happening? I am on medication but still in pain and getting more complicated illnesses…. Why chronic diseases? May be each starts on its own, maybe they are not even related…*,” Malawi IDI PLWMM Female 40-49 (HIV, HTN).

Living with multimorbidity profoundly disrupted daily life, affecting physical health, emotional wellbeing, and social roles. The progressive accumulation of chronic conditions lead to reduced independence, financial strain, and heightened emotional distress. For many, stigma and limited awareness worsened the experience, particularly when visible symptoms or disabilities result in diminished sense of family or community belonging.

### Self-management and adaptation

PLWMM and carers described their experiences of self-management as the combination of complex interactions regarding physical, medical, emotional and diet management. Despite the demonstrated willingness to engage with biomedical treatments especially as the number of comorbidities increased, PLWMM reported significant barriers to access, including frequent drug shortages and high cost of drugs. High out of pocket (OOP) expenses were particularly troublesome for participants. Both settings had shortages of DM and CKD drugs with ARTs being the most readily available treatment.

> *“Diabetes terrifies me… I can’t live without insulin so it should always be available. But often there are no drugs at the hospital and mother doesn’t have money, she’s not making enough sales, so we must buy, and it’s expensive…. often, she borrows money,”* Malawi IDI PLWMM Male, 20-29 (HIV, HTN, DM).
>
> *“I explained my financial situation, but they advised me to ask my children for the money. I expressed my concern, given that they had encouraged people to seek medical care. I left without obtaining the medication,”* Tanzania, IDI PLWMM Female 60-69 (DM, HTN).

While catastrophic expenditure persisted among patients with CKD multimorbidity across both settings, Tanzania reported increased personal expenditure compared to Malawi. Although patients with CKD multimorbidity in Tanzania seemingly had improved initial access to treatment than in Malawi, continuity of care was a challenge. Most PLWMM with CKD reported discontinuation of treatment due to catastrophic expenditures.

> *“At KCMC I was told to go to the kidney clinic, I don’t even have a hundred shillings, I didn’t go until today. aah! I only use pressure washers. They told me to go back to the clinic, but I didn’t,… I didn’t have any money, even my mother doesn’t…, she sold all her cows until they were all gone,”* Tanzania, IDI PLWMM Female 30-39 (HTN, CKD)

Multimorbidity self-management frequently included medical pluralism such as herbal or traditional medicine with the practice more commonly acknowledged in Tanzania. In Malawi, participants seemed reluctant to disclose herbal use, possibly due to fear of judgment from researchers or health workers. Medical pluralism varied based on the number or combination of conditions and trust in the effectiveness of combined treatments. People living with HIV as a comorbidity were the least likely to use herbal treatments, as they reported having adequate access to information and biomedical care and felt their condition was well controlled. Individuals with CKD comorbidity were also less inclined to use herbal treatments due to concerns about potential complications, particularly the risks associated with drug interactions.

> *“The challenge is that there’s lots of ‘street doctors’, … it’s confusing, sometimes you believe and follow such advice. He suffered because we used herbs, he got worse, so we decided to stop, and follow hospital treatment only,”* Tanzania 30-39 FGD Carer Female.
>
> *“Before kidney transplant, I would use traditional medicine, but I stopped after the transplant because I am afraid that I may die. One of my friends died from it,”* Tanzania IDI PLWMM Male 60-69 (DM, HTN, CKD),

Across both settings, it was common for PLWMM to move away from using traditional or herbal treatments as the number of conditions increased or worsened. This shift signified reduced trust in the effectiveness of herbal treatments as conditions progressed over time. Some participants adopted exclusive biomedical treatment.

> *“I have passed through them, but now, I can say I have given up about mere people or traditional healer because my conditions keep deteriorating,”* Malawi, IDI PLWMM Male 40-49 (HTN, DM).
>
> *“The doctor advised me to stick to a recommended diet…. I used numerous herbs but eventually stopped because despite using a diet and herbs, the situation was deteriorating. In the end, I only used hospital medication,”* Tanzania FGD Female PLWMM 20-29 (DM, HTN).

A few PLWMM and their carers considered herbs as non-toxic and safer than biomedical treatments, which they associated with chemicals and long-term side effects.

> *“Traditional remedies like herbs are not as toxic as the hospital ones, that’s why some people use them,”* Tanzania FGD Carer (Patient 50-59 CKD, HTN).

PLWMM and carers often identified specific moments/events in their disease progression which they believed influenced key decision-making regarding self-management. In both settings, such tipping points were more commonly noted among patients diagnosed with DM or HTN. Examples of such defining events would be the temporal discontinuation of drugs or a social event such as a disagreement with a loved one.

> *“I was fine until I started arguing with my wife, I started overthinking, wondering if I should divorce her then I fell sick and was diagnosed with BP…. From nowhere I was diagnosed with diabetes,”* Malawi FGD PLWMM Male 40-49 (HIV, HTN, DM).
>
> *“It’s okay with HIV, I forget it because of the drugs that I take but diabetes, once I didn’t take medication for two days because it was not available, I experience serious symptoms. I am facing the symptoms to this day,”* Tanzania IDI PLWMM Woman 60-69 (HIV, DM, HTN).

Further, PLWMM and their carers experienced emotional stress which affected how they self-managed. Emotional stress related to pain, social isolation and lack of essentials to manage their conditions. In Tanzania, older PLWMM and younger women in Malawi were more likely to acknowledge emotional stress.

> *“I have been having these challenges since my children moved out - no one sends money or food. I have no food, no energy. Every day, I must look for food. What will happen when I am not able to do casual work just to earn a little like K500?”* Malawi IDI PLWMM Female 40-49 (HIV, HTN).

Across both settings, self-management decisions among PLWMM were shaped by individual-level constraints such as financial limitations and limited health literacy. Faced with persistent challenges, participants often prioritised one condition over others, guided by perceived severity; symptom urgency; impact on daily functioning; and practicality of care. CKD was most often prioritised, followed by DM over HTN. HIV was typically deprioritised, as PLWMM had regular access to ART and understood that consistent adherence led to viral suppression. While many aimed for holistic self-management, structural and personal barriers often forced a focus on the most immediate or life-threatening concern.

> *“Sometimes the food isn’t enough, when I consider this with the strength of the medication, I would not inject myself. Maybe that’s why the condition worsened. Sometimes we eat whatever is available and this could have raised the sugar levels,”* Malawi IDI PLWMM Male 40-49 (DM, HTN, CKD).
>
> *“If you don’t have insulin, you can die with diabetes, so you have to get diabetes medication. The rest should follow because, without insulin, life has stood still, so I look for the priority,”* Tanzania FGD PLWMM Male 50-59 (DM, HTN).

In response to pain, fatigue and limited mobility, PLWMM and carers in both settings integrated light physical activity, for example routine chores.

> *“I make her do light chores like cutting vegetables, sweeping,”* Tanzania FGD Carer Male 50-59.

While family and community support were pivotal to self-management, female participants more often engaged in peer-led support groups that facilitated self-care, emotional support, and health information sharing. These groups enabled members to monitor BP and sugar levels, record results, and advise one another. However, these networks focussed on one condition rather than multimorbidity, in line with healthcare provision with HIV discussed at ART clinics, whilst DM and HTN at a separate clinic.

> *“We have a BP and sugar support group, there are 5 of us who volunteer, so when we go to the hospital on clinic day…, we test sugar by ourselves, then we document in the health passport book, we advise each other,”* Malawi FGD PLWMM, Female 40-49 (HTN and DM).

Across both Malawi and Tanzania, self-management among PLWMM and their carers was multifaceted, with biomedical care being highly valued, especially as conditions worsened or accumulated. Many participants identified tipping points along their illness journey when multimorbidity either improved or deteriorated. CKD and DM posed the greatest burden due to their impact, high out of pocket costs, and limited treatment availability. Concerns about side effects often influenced decisions on using traditional care, highlighting the complexities of medical pluralism.

### Prioritisation of individual diseases within the multimorbidity paradigm

PLWMM with HTN often reported temporary treatment discontinuation due to concerns about lifelong medication; limited understanding of the necessity for ongoing treatment; beliefs regarding age-related risk; and/or lack of drugs. In the absence of overt hypertensive symptoms, patients expressed apprehension about potential side effects of long-term medication. Limited awareness, overt symptoms and a lack of medical recommendations led some PLWMM to adopt alternative strategies, such as lifestyle changes, instead of using these as complementary approaches. This behaviour highlighted a gap in understanding of HTN as a chronic condition, and the necessity of maintaining consistent, long-term medication adherence for effective disease management.

> *“Mostly, they notice high blood pressure when I have malaria. They inquire if I am on treatment and prescribe antihypertensive medication. I would take until my BP normalises because it becomes elevated when I have malaria…, after improvement, I would stop. … I didn’t want to take the drugs every day, I preferred exercises, so often, my BP would be normal,”* Malawi IDI PLWMM Male 50-59 (HIV, HTN).

Age-related concerns about initiation of HTN treatment often centered around the potential long-term side effects of medications. Treatment discontinuation was both health worker- and patient-initiated and common in younger patients (aged ≤40) in both settings. In Malawi, patients described being advised to monitor their blood pressure, though this was often passive and lacked a clear structure or follow-up plan. Age-related concerns in Malawi may be reflective that younger participants tended to be recruited during this study (median age of 50 years [IQR: 44–57; range: 26–69] for PLWMM in Malawi, versus 60 years [IQR: 47–66; range: 31–80] in Tanzania).

> *“They said my blood pressure was under control and considering my age, they said ‘it’s not good that I should start taking blood pressures medication now* (2010). *So maybe we need to wait, and we will be monitoring you. You should keep coming for check-ups and if we notice that its high, then we will restart the treatment’,”* Malawi IDI PLWMM Female 40-49 (HIV, HTN).

While patient reasons for discontinuation included fear of lifelong treatment and limited health literacy, it was often unclear what had been discussed with health workers, making it difficult to determine how treatment plans had been developed (discontinuation is not typically recommended in normative clinical guidelines (44)). Some PLWMM who had their HTN medications discontinued, suffered secondary complications of uncontrolled disease. Some described experiencing stroke-like symptoms while others required another hospital admission. While these experiences sometimes reflected patient engagement with their conditions, they also underscore broader challenges related to quality of care including drug stockouts and poor continuity in the management of chronic conditions. Although the attribution of DM onset to interrupted HTN treatment may not be medically accurate, it illustrates the consequences of limited health literacy, a phenomenon that is not solely a patient-level issue, but one that reflects systemic failures in communication, counselling, and coordinated care. Treatment discontinuation was especially common among those with HTN.

> *“Once there was a shortage of hypertension drugs at the hospital and after some time, we noted the patient would tired, weak, and shiver. When we visited the hospital, he was diagnosed with diabetes, I think was a result of the interruptions to his anti-hypertension treatment,”* Malawi FGD Female Carer 50-59.
>
> *“Regarding HIV, I don’t think there is much concerning food, I cook to be full and just take medication… But to control hypertension, I reduce like the salt*.,*”* Malawi PLWMM iDI 60-69 (HIV, DM).

## Discussion

We found PLWMM and carers in Malawi and Tanzania face distress due to reduced physical functionality and diminished sense of self-identity, exacerbated by financial and social pressures, and limited access to healthcare. This is important because engaged and empowered patients are key to successful management of chronic conditions. Further work is required to understand how our findings are generalisable in other Southern African contexts. We recommend increased health education to improve health literacy; peer-led interventions; and collaborative goal setting between healthcare providers, PLWMM, and their carers to reduce structural barriers and augment community-based support systems.

A key aim of our work was to explore lived experiences of PLWMM and their carers and to explore the relevance of an expanded conceptual model for multimorbidity in Africa (6). We have demonstrated the dynamic interplay of biomedical, social, and economic factors that shape how PLWMM and their carers experience multimorbidity. While biomedical care often focuses on chronic conditions, PLWMM frequently prioritise acute symptoms; financial hardship; stigma; and shifting social roles. Self-management of multimorbidity reflects a continuous negotiation shaped by disease combinations, age, gender, and context. These complex factors strongly shape patient understanding and response to multimorbidity, more so than individual disease labels. In addition, our findings demonstrated contextual nuances with a dynamic interaction between biomedical and social factors (12, 40, 45). For instance, HIV was frequently perceived as the least distressing comorbidity; younger men more commonly internalised stigma; women often prioritised caregiving over their own needs; and older adults with DM and HTN reported emotional stress linked to dependency and lost independence. In Malawi, younger PLWMM expressed particular concerns about initiation of lifelong antihypertensive treatment, often citing fears of long-term dependence and side effects. Across both settings, those living with CKD comorbidity experienced the greatest treatment burden, discontinuation of care due to catastrophic OOP expenditure was reported more often in Tanzania.

These varied narratives emphasise the need for person-centred care that responds to individual con-texts, illness pathways, and evolving priorities. Further to the individuality of perceptions, the attribution of conditions with visibly apparent symptoms to spiritual or moral causes particularly in Malawi reflects how illness is often interpreted beyond biomedical reasoning. These interpretations, rooted in social and cultural beliefs, linked illness to wrongdoing, misfortune, or supernatural causes such as witchcraft, reinforcing blame and exclusion, and intensifying emotional distress and marginalisation, phenomena also reported in South Africa (46, 47) and Ethiopia (48).

The concept of “tipping points” described in the Expanded Conceptual Model of Multimorbidity (6) was evident in our data, underlining the need for responsive, adaptable care strategies. Participant descriptions of temporary disruptions such as missing drugs due to medication stockouts or emotional distress setting off a cascade of events that disrupted self-management and worsened their conditions highlight gaps in health literacy amongst participants and broader structural issues. Limited health systems support further exacerbated reactive self-management amongst participants and fragmented care for NCDs (49-51). Tailoring services through more flexible scheduling, extended consultation times, and integrated peer support could support care delivery for the complex, evolving needs of people living with multimorbidity (52). Peer-led interventions, co-designed with PLWMM, could draw on lessons from HIV programmes. Evidence from Rwanda suggests such peer support can improve mental health, quality of life, and social inclusion among people with complex needs (53).

As self-management extends beyond individual with disease, leveraging informal support systems can build collective capacity, especially where formal care is limited. Collaborative goal setting can help define realistic, context-appropriate objectives that foster self-confidence and promote sustainable habits. This approach can foster keystone habits, strengthen self-confidence, and support more meaningful, sus-tained management of multimorbidity (12, 54). Additionally, integration of low-cost self-monitoring tools such as paper-based tracking systems, pill counting, and peer monitoring can empower patients to track symptoms, medication adherence, and treatment progress (55). While existing literature on single-disease self-monitoring shows promise (55, 56), the relationship between self-efficacy and willingness to self-monitor among PLWMM remains underexplored.

Primary and secondary prevention are critical for delaying onset, accumulation or complication of multimorbidity. In Malawi, integration of chronic care into primary-level services through community outreach and task-shifting has improved early detection and continuity of care (57). Community-based NCD screening, when aligned with existing programmes, is also feasible and effective (58). Among participants in both Malawi and Tanzania, experiences of living with multimorbidity were shaped by individual-level barriers. These include financial hardship and limited health literacy, particularly around DM, HTN, and CKD and systemic factors, including lack of educational support; medication stockouts; and poorquality health worker-patient interactions. Strengthening primary health facilities and expansion of community outreach may help bring care closer to home, enabling earlier diagnosis, improved continuity of care, and reduced complications associated with multimorbidity (57). Further research could explore effectiveness of these approaches in the context of multimorbidity. These findings support calls to strengthen and implement the basic health packages in both Malawi and Tanzania (12, 59, 60).

While the Expanded Model of Multimorbidity is primarily designed to support understanding of lived experiences from multidisciplinary perspectives, there is an expectation that such frameworks could also inform prevention strategies and strengthen the role of community-based care. However, the model gives limited attention to these aspects, which are particularly critical in contexts like Malawi and Tanzania. While this is a key step, the model could also help guide context-specific research and interventions that address the needs of PLWMM in resource-limited settings.

### Strengths and limitations

A key strength of the study was its prolonged engagement with participants, which facilitated a deep understanding of patient and carer experiences with multimorbidity. The comparative design enabled identification of cross-site patterns and divergences, enriching analysis by showing how contextual factors shaped lived experiences and care practices. Triangulation of methods further strengthened the validity of findings. Furthermore, our application of a framework grounded in multiple validated theories enhances the robustness and confidence in our findings. Whilst we aimed to balance PLWMM and carers by site and disease combination using purposive sampling, this was impractical due to the variability in patient presentations. Consequently, we recruited patients as they presented for care rather than through a matched approach. Additionally, we did not specifically include or exclude patients with depression as part of our criteria. Given the high prevalence of depression among patients with chronic conditions in LMIC, more intentional inclusion could have provided valuable insights into the results. Participants may have also downplayed their use of informal care due to our biomedical focus. However, with limited access to medical care, combining treatments is likely common. Future research should explore how people manage multimorbidity with multiple therapies. Coding by one researcher may introduce bias or limit interpretive breadth; however, consistency and deep data familiarity were strengths. Although we recruited from both rural and urban sites, participants frequently sought care across both settings depending on perceived needs and availability of transport money, making it difficult to draw meaningful rural-urban comparisons. Lastly, study recruitment predated recent USAID HIV funding cuts, which may affect care continuity and the relevance of some findings over time (61).

## Conclusions

Living with multimorbidity in Malawi and Tanzania requires navigation of complex biomedical, social, and systemic challenges. Person-centred, integrated care must respond to these intersecting realities. Co-creation of care pathways with PLWMM and carers through effective communication and collaborative goal setting is key. Health education on disease interactions can support informed self-care, while reducing structural barriers and strengthening primary-level NCD services are essential for sustained management. Peer support networks offer vital emotional and practical help, reinforcing community-based care and reducing isolation. Although men did not report similar participation, this suggests a gap in access or acceptability. Future interventions should explore gender-sensitive ways to engage men in peer support aligned with their needs and norms.

## Acknowledgements

The lead author would like to express our gratitude to patients and carers in Malawi and Tanzania who shared with us information about their personal lives and experiences.

## Author contributions

### Conceptualisation

Sangwani N. Salimu, Ben Morton, Deborah Nyirenda, Nicola Desmond

### Data collection

Sangwani N. Salimu, Alice Rutta, Martha Oshoseny, Graciana Kimario, Ibrahim Simiyu

### Formal analysis

Sangwani N. Salimu

### Funding acquisition

Jamie Rylance, Ben Morton, Adamson Muula, Augustine Choko, Miriam Taegtmeyer, Matthew Rubach, Charity Salima, Mulinda Nyirenda, Sarah White, Paul Dark, Felix Limbani, Eve Worrall

### Supervision

Ben Morton, Nicola Desmond, Deborah Nyirenda

### Writing – original draft

Sangwani N. Salimu

### Review & editing

Stephen A. Spencer, Alice Rutta, Treighcy Gift Banda, Martha Oshoseny, Naiteya Yongolo, Marlen Chawani, Augustine Choko, Julian T. Hertz, Gimbo Hyuha, Blandina T. Mmbaga, Juma Mfinanga, Rhona Mijumbi, Francis Sakita, Charity Salima, Hendry Sawe, Ibrahim Simiyu, Miriam Taegtmeyer, Sarah Urasa, Nateiya M. Yongolo, Adamson Muula, Charity Salima, Mulinda Nyirenda, Paul Dark, Jamie Rylance, Eve Worrall, Felix Limbani, Nicola Desmond, Deborah Nyirenda, Ben Morton.

## Data availability statement

The data set used and analysed during this study is available from the corresponding author on reasonable request. Please contact Sangwani N. Salimu.

## Supplementary Information

**Table S1.** Malawi- Demographic characteristics of PLWMM involved in IDIs.

Malawi- Demographic characteristics of PLWMM involved in IDIs
| Participant ID | Sex | Age range | Conditions | Other conditions |
| --- | --- | --- | --- | --- |
| 1 | Female | 31-50 | HIV, Hypertension | TB, Meningitis, Appendicitis |
| 2 | Male | 31-50 | HIV, Hypertension | TB |
| 3 | Male | 30 and below | HIV, Hypertension | Heart failure |
| 4 | Female | 31-50 | HIV, Hypertension | Heart Failure |
| 5 | Female | 51-70 | HIV, Hypertension, Diabetes | Heart failure |
| 6 | Female | 31-50 | HIV, Hypertension, Diabetes | Heart Failure |
| 7 | Male | 51-70 | HIV, Hypertension | Heart failure |
| 8 | Male | 30 and below | HIV, Hypertension, Diabetes |  |
| 9 | Male | 51-70 | Hypertension, Diabetes | Pneumonia |
| 10 | Male | 31-50 | Hypertension, Diabetes | Pancreatitis |
| 11 | Female | 51-70 | HIV, Hypertension | TB |
| 12 | Female | 31-50 | HIV, CKD |  |
| 13 | Female | 51-70 | Hypertension, Diabetes |  |
| 14 | Male | 31-50 | Hypertension, Diabetes, CKD |  |
| 15 | Female | 51-70 | Hypertension, Diabetes |  |
| 16 | Male | 51-70 | Hypertension, HIV |  |

**Table S2.** Tanzania- Demographic characteristics of PLWMM involved in IDIs.

| **Participant ID** | **Sex** | **Age** | **Condition** | **Other conditions** |
| --- | --- | --- | --- | --- |
| 1 | Female | 51-70 | Hypertension, Diabetes | Urinary Track Infection |
| 2 | Female | 31-50 | HIV, Diabetes | TB |
| 3 | Male | 71 and above | Hypertension, Diabetes |  |
| 4 | Male | 51-70 | Hypertension, Diabetes |  |
| 5 | Female | 51-70 | Hypertension, Diabetes | Stroke |
| 6 | Female | 51-70 | Hypertension, Diabetes |  |
| 7 | Female | 31-50 | Hypertension, CKD |  |
| 8 | Female | 31-50 | Hypertension | TB, Liver failure |
| 9 | Male | 31-50 | Hypertension, Diabetes | TB, Heart Failure |
| 10 | Male | 51-70 | Hypertension, Diabetes |  |
| 11 | Male | 51-70 | Hypertension, Diabetes | Stroke |
| 12 | Male | 31-50 | Hypertension, CKD |  |
| 13 | Male | 51-70 | Hypertension, CKD |  |
| 14 | Female | 51-70 | Hypertension, CKD |  |
| 15 | Female | 51-70 | Hypertension, Diabetes | Vertebral Disk Condition |
| 16 | Male | 51-70 | Hypertension Diabetes CKD |  |
| 17 | Male | 51-70 | Hypertension, Diabetes, CKD |  |
| 18 | Male | 51-70 | Hypertension, Diabetes, CKD | TB |
| 19 | Female | 31-50 | Hypertension, CKD |  |
| 20 | Male | 31-50 | Hypertension, CKD |  |
| 21 | Female | 51-70 | Hypertension, Diabetes | Stroke |

